# Cardiac Rehabilitation Participation in Heart Failure Over Time: An Analysis of the Colorado All Payer Claims Database

**DOI:** 10.1101/2023.03.01.23286662

**Authors:** Lubin R. Deng, Vincenzo B. Polsinelli, Vinay Kini, Morgan Wilson, Pamela Peterson, David B. Bekelman, Kelsey M. Flint

**Author notes:** **Corresponding Author:** Vincenzo B. Polsinelli, MD, Fellow, Cardiovascular Disease, University of Colorado, 12605 East 16th Avenue, 3rd Floor, Aurora, CO 80045.

## Abstract

**Background:** Despite CMS coverage expansion supporting cardiac rehabilitation (CR) in patients with heart failure (HF) in 2014, data suggest that utilization among patients with HF is low. We describe CR participation and adherence among HF patients in Colorado.

**Methods:** Data from the Colorado All-Payer Claims Database from 2010-2018 were used. Patients with HF were identified by ≥2 claims with a HF diagnosis code, then grouped by type of HF (HFrEF, HFpEF, unspecified). CR participation and adherence were identified using CR CPT codes. Participation rates were calculated by quarter of each year. Cochran-Armitage tests determined whether temporal trends were significant. Association between CR participation and payer source was examined in adjusted logistic regression models.

**Results:** 263,476 patients with HF were identified. 4.77% of all HF patients attended CR at least once; this result was similar for HFpEF (4.35%), unspecified HF (4.15%), and higher in the HFrEF group (8.25%). Overall adherence was poor (median 8 visits, IQR 3-18; full adherence=36 visits). CR participation over time increased (P<0.01) for all HF patients. Compared to patients with commercial insurance, patients with Medicare, Medicaid, or Medicare Advantage were less likely to have participated in CR at least once (P<0.01). Race, sex and presence of another indication for CR were also associated with at least one CR visit (P<0.01).

**Conclusions:** In the state of Colorado, CR participation improved from 2010-2018 among all patients with HF. Our data suggest that payer source, race, sex and presence of another indication for CR drive CR participation in patients with HF.

## Introduction

Cardiac rehabilitation (CR) is a comprehensive lifestyle program that includes moderate intensity aerobic exercise for 36 supervised exercise sessions over 12 weeks.^1-3^ Evidence suggests that CR can improve functional capacity, quality of life, and reduce mortality and hospitalization for patients with heart failure (HF), with the most robust data for those with heart failure with reduced ejection fraction (HFrEF).^3-7^ Although regular exercise has been a Class I recommendation for patients with HF since 2005,^8^ CR has only been a guideline recommendation for patients with heart failure since 2013, but was also restated in the 2022 guidelines.^4^ [Heidenrich] (working on end note)

However, use of CR among patients with heart failure is particularly low. A study of patients hospitalized for heart failure between 2005 and 2014 found that just 12% of patients with HFrEF were referred to CR on hospital discharge.^9^ Other studies have indicated that the CR referral rates of patients with heart failure are lower than the referral rates of patients with other indications for CR.^10,11^

There are many documented barriers to CR participation in patients with HF. Historically, lack of insurance coverage was the most significant barrier; however in February 2014, the Centers for Medicare and Medicaid Services (CMS) announced coverage for CR participation in patients with HFrEF (EF ≤ 35%),^12^ representing the first move to pay for CR in a HF population.^13^ Studies predicted that CR participation in patients with HF would increase after 2014 due to expanded coverage alongside other active efforts, such as computerized prompts for referral and providing early CR appointments after discharge.^13-16^ In the few years following the 2014 CMS coverage expansion, it became increasingly apparent that these efforts were insufficient for significantly augmenting CR participation among patients with HF.^17^ However it is unknown whether this finding holds true across a longer time period, whether there are differences in CR participation and adherence by type of HF (HFrEF vs. HFpEF), presence of non-HF indication for CR or by payer source (commercial, Medicare, Medicaid, Medicare Advantage).

In this study, we examined CR participation and adherence rates in all patients with HF in the state of Colorado between 2010 and 2018, by type of HF, presence of non-HF indication for CR and payer source. We hypothesized that participation in CR would be very low but would increase by a statistically significant amount over time in a) patients with HFrEF, and b) all patients with HF who also had a non-HF indication for CR.

## Methods

### Data source

We used data from the Colorado All-Payer Claims Database (APCD), which is a comprehensive administrative dataset that includes inpatient, outpatient, physician, and facility claims on nearly all patients who received care in the state. Reporting is required for all insurance companies and plans with the exception of federal health facilities (e.g., Veterans Health Administration hospitals) and self-insured group health plans. The APCD also includes beneficiary demographics including age and sex, insurance carrier, and hospital identifiers, but does not include reliable data on race/ethnicity. Data from January 2010 to December 2018 were available for this study. Both inpatient and outpatient claims were included.

### Defining the cohort

Patients with heart failure were identified using the criterion of having at least two claims with a HF diagnosis code (ICD-9 code 428 or ICD-10 code I50). These patients were divided into groups by type of HF: HFrEF (ICD-9 code 428.2X or 428.4X; ICD-10 code I50.2X, I50.4X, or I50.82), HFpEF (ICD-9 code 428.3X; ICD-10 code I50.3X), unspecified (ICD-9 code 428.9, 428.0, 428.1, or 428; ICD-10 code I50.9, I50.1, I50.83, or I50.89), or right HF (ICD-10 code I50.81, I50.810, I50.811, I50.812, I50.813, or I50.814). Patients with end-stage HF (ICD-10 code I50.84) were excluded.

### Definition of the primary exposure variable

Cardiac rehabilitation visits were identified using CPT codes of 93797 and 93798. CR participation rate was calculated for each group of patients by quarter of each calendar year from 2010 to 2018. For each group, the numerator for each quarter was the number of patients who had at least one CR visit in that quarter (may repeat from year to year, but each patient only counted once per four quarters). The denominator for each quarter was the number of patients with a claim for the relevant type of HF in that quarter. Patients who participated in CR were then removed from the denominator for the following 3 quarters.

### Definition of Covariates

For each type of HF, patients were further grouped by whether they had an additional indication for cardiac rehabilitation: coronary artery disease (ICD-9 code 414; ICD-10 code I25), history of surgical valve replacement or transcatheter aortic valve replacement/implantation (ICD-9 code V42.2 or V43.3; ICD-10 code Z95.2, Z95.3, or Z95.4), history of CABG (ICD-9 code V45.81; ICD-10 code Z95.1), history of PCI (ICD-9 code V45.82; ICD-10 code Z95.5), claudication (ICD-9 code 443.9 or 440.21; ICD-10 code I73.9, I70.21, I70.31, I70.41, I70.51, I70.61, or I70.71), angina (ICD-9 code 413; ICD-10 code I20, I25.7, or I25.11), left ventricular assist device (ICD-9 code V43.21; ICD-10 code Z95.811), or heart transplant (ICD-9 code V42.1; ICD-10 code Z94.1).

Other comorbid conditions were identified using the following diagnostic codes: atrial fibrillation (ICD-9 code 427.31; ICD-10 codes I48.0, I48.1, I48.2, I48.91), hypertension (ICD-9 code 401; ICD-10 code I10), hyperlipidemia (ICD-9 codes 272.0, 272.1, 272.2, 272.3, 272.4; ICD-10 codes E78.0, E78.1, E78.2, E78.3, E78.4, E78.5), peripheral vascular disease (ICD-9 code 443; ICD-10 code I73), tobacco use (ICD-9 code V15.82; ICD-10 code Z72.0), chronic obstructive pulmonary disease (ICD-9 codes 490, 491, 492, 493, 494, 495, 496; ICD-10 code J44), cancer (ICD-9 codes 140-209; ICD-10 codes C00-C96), chronic kidney disease (ICD-9 code 585; ICD-10 code N18), diabetes mellitus (ICD-9 code 250; ICD-10 codes E08, E09, E10, E11, E13).

### Statistical Analysis

Data are presented as counts, percentages, means, standard deviations, medians, and interquartile ranges, depending on the circumstances. Presence of a temporal trend in CR participation rates from the first quarter of 2010 to the fourth quarter of 2018 was tested for every group using the Cochran-Armitage test at a significance level of 0.05.

We examined whether payer source (commercial, Medicare, Medicaid, Medicare Advantage) was a predictor of CR participation using logistic regression. Demographic factors (age, sex race), type of HF (HFrEF, HFpEF, unspecified), presence of at least one additional indication for CR, and individual comorbidities (coronary artery disease, history of valve procedure [surgery or transcatheter aortic valve replacement], atrial fibrillation, hypertension, hyperlipidemia, peripheral vascular disease, tobacco use, chronic obstructive pulmonary disease, cancer, chronic kidney disease, and diabetes mellitus) were included as covariates. The primary analysis utilized the whole sample of patients with HF. Sensitivity analyses were performed in the following cohorts: only patients with HFrEF, and only in the years since the Medicare coverage expansion for CR for HFrEF (2014-2018).

## Results

A total of 264,073 patients with heart failure were included in this study. Descriptive characteristics of the participants at baseline are provided in **Table 1**. The participants had a mean age of 74.3±16.2 years, were predominantly white (75.7%), 52.7% were female, and most were enrolled in Medicare (55.69%). Comorbid conditions that are also indications for CR were prevalent among the participants (e.g., 57.0% had coronary artery disease), and cardiac and non-cardiac comorbidities were common as well. (**Table 1**)

**Table 1:** Characteristics of Target Population (N = 263,476)

| Characteristic, n (%) | Overall<br>(N=263,476) | HFrEF<br>(n=38,127) | HFpEF<br>(n=41,164) | Unspecified<br>(n=184,185) | Number<br>Missing |
| --- | --- | --- | --- | --- | --- |
| **Demographics:** |  |  |  |  |  |
| Age, years, mean (SD) | 74.3 (16.2) | 70.9 (16.3) | 73.4 (13.4) | 75.3 (16.5) | 7,059 |
| Race: |  |  |  |  | 100,889 |
| American Indian/Alaska Native | 3,360<br>(2.07%) | 438<br>(2.11%) | 323<br>(1.36%) | 2,599<br>(2.19%) |  |
| Asian | 8,465<br>(5.21%) | 790<br>(3.80%) | 672<br>(2.84%) | 7,003<br>(5.91%) |  |
| Black | 6,995<br>(4.30%) | 1,016<br>(4.89%) | 911<br>(3.85%) | 5,068<br>(4.28%) |  |
| Native Hawaiian or other Pacific Islander | 132 (0.08%) | 22 (0.11%) | 6 (0.03%) | 104 (0.09%) |  |
| White | 123,362<br>(75.87%) | 15,138<br>(72.84%) | 18,669<br>(78.81%) | 89,555<br>(75.59%) |  |
| Other | 20,637<br>(12.69%) | 3,378<br>(16.25%) | 3,109<br>(13.12%) | 14,150<br>(11.94%) |  |
| Female gender | 135,098<br>(52.72%) | 14,777<br>(40.3%) | 22,548<br>(57.71%) | 97,771<br>(54.1%) | 7,225 |
| **Insurance type:** |  |  |  |  | 443 |
| Medicare | 146,727<br>(55.78%) | 17,246<br>(45.32%) | 20,912<br>(50.88%) | 108,569<br>(59.05%) |  |
| Medicare Advantage/Managed Medicare | 54,446<br>(20.70%) | 10,052<br>(26.41%) | 10,935<br>(26.60%) | 33,459<br>(18.20%) |  |
| Medicaid | 40,450<br>(15.38%) | 6,226<br>(16.36%) | 5,073<br>(12.34%) | 29,151<br>(15.85%) |  |
| Commercial | 21,410<br>(8.14%) | 4,532<br>(11.91%) | 4,182<br>(10.17%) | 12,696<br>(6.90%) |  |
| **Additional indications for cardiac rehabilitation:** |  |  |  |  |  |
| Coronary artery disease | 149,775<br>(57.09%) | 25,780<br>(67.9%) | 20,701<br>(50.5%) | 103,294<br>(56.3%) | 930 |
| History of surgical valve replacement | 14,817<br>(5.65%) | 2,479<br>(6.55%) | 2,263<br>(5.53%) | 10,075<br>(5.49%) | 1,213 |
| History of transcatheter aortic valve replacement/implantation | 11,117<br>(4.24%) | 1,631<br>(4.31%) | 1,376<br>(3.36%) | 8,110<br>(4.42%) | 1,243 |
| History of CABG | 19,790<br>(7.54%) | 4,881<br>(12.89%) | 3,027<br>(7.39%) | 11,882<br>(6.48%) | 1,173 |
| History of PCI | 31,663<br>(12.07%) | 6,782<br>(17.90%) | 4,186<br>(10.22%) | 20,695<br>(11.28%) | 1,147 |
| Claudication | 63,885<br>(24.35%) | 7,993<br>(21.10%) | 9,498<br>(23.19%) | 46,394<br>(25.28%) | 1,125 |
| Angina | 31,813<br>(12.13%) | 6,449<br>(17.02%) | 4,833<br>(11.80%) | 20,531<br>(11.19%) | 1,148 |
| Left ventricular assist device | 216 (0.08%) | 54 (0.14%) | 5 (0.01%) | 157 (0.09%) | 1,263 |
| Heart transplant | 629 (0.24%) | 129<br>(0.34%) | 49 (0.12%) | 451 (0.25%) | 1,263 |
| **Cardiac comorbidities:** |  |  |  |  |  |
| Atrial fibrillation | 112,149<br>(42.72%) | 17,490<br>(46.11%) | 15,482<br>(37.77%) | 79,177<br>(43.12%) | 933 |
| Hypertension | 230,105<br>(87.65%) | 32,743<br>(86.22%) | 36,474<br>(88.86%) | 160,888<br>(87.53%) | 657 |
| Hyperlipidemia/<br>dyslipidemia | 181,584<br>(69.17%) | 27,057<br>(71.24%) | 30,448<br>(74.18%) | 124,079<br>(67.52%) | 691 |
| Peripheral vascular disease | 62,963<br>(23.98%) | 7,802<br>(20.60%) | 9,199<br>(22.47%) | 45,962<br>(25.04%) | 1140 |
| Coronary artery disease | 149,775<br>(57.09%) | 25,780<br>(67.9%) | 20,701<br>(50.5%) | 103,294<br>(56.3%) | 930 |
| Tobacco use | 44,180<br>(16.83%) | 6,866<br>(18.13%) | 5,866<br>(14.33%) | 31,448<br>(17.14%) | 1192 |
| **Non-cardiac comorbidities:** |  |  |  |  |  |
| Chronic obstructive pulmonary disease | 136,697<br>(52.07%) | 17,325<br>(45.70%) | 19,867<br>(48.47%) | 99,505<br>(54.19%) | 955 |
| Cancer (breast, prostate, colorectal, lung) | 41,916<br>(15.97%) | 5,361<br>(14.15%) | 6,505<br>(15.89%) | 30,050<br>(16.38%) | 1153 |
| Chronic kidney disease | 158,945<br>(60.54%) | 23,408<br>(61.69%) | 24,361<br>(59.39%) | 111,176<br>(60.51%) | 790 |
| Diabetes mellitus | 111,514<br>(42.48%) | 15,602<br>(41.16%) | 16,766<br>(40.92%) | 79,155<br>(43.11%) | 998 |

Cardiac rehabilitation participation rates were very low for all HF subgroups (**Table 2**). Only 4.77% of participants attended cardiac rehabilitation at least once, and this percentage was highest among patients with HFrEF (8.25%). Continuation and completion rates for CR programs were also poor. The patients who did participate in CR attended a median of only 8 out of 36 visits in total. Only 1.24% of participants attended at least half (18) of the visits, and 0.58% (1.06% for HFrEF) completed all 36 CR visits.

**Table 2:** Cardiac Rehabilitation Participation by Type of HF

|  | **Overall** | **HFrEF** | **HFpEF** | **Unspecified** |
| --- | --- | --- | --- | --- |
| Number (%) of patients with at least one cardiac rehabilitation visit | 12,590<br>(4.77%) | 3,146<br>(8.25%) | 1,790<br>(4.35%) | 7,645<br>(4.15%) |
| Of those who participated in cardiac rehabilitation, median (IQR) number of visits | 8 (3-18) | 9 (4-21) | 8 (4-20) | 8 (3-17) |
| Number (%) of patients who attended 50% (18) or more visits | 3,263<br>(1.24%) | 895<br>(2.35%) | 499<br>(1.21%) | 1,866<br>(1.01%) |
| Number (%) of patients who completed all 36 visits | 1,538<br>(0.58%) | 406<br>(1.06%) | 240<br>(0.58%) | 891 (0.48%) |

Trends of CR participation rates from 2010 to 2018 are illustrated in **figure 1**, however several findings are worth highlighting. Participation rates increased from 0.51% in Q1 of 2010 to 1.09% in Q4 of 2018 in all patients with HF (P<0.01). In HFrEF, participation increased from 0.66% to 1.99% (P<0.01), and from 0.53% to 1.04% in HFpEF (P<0.01). CR participation rates also increased among those who had had additional indications for CR (1.52% in Q1 of 2010 to 2.68% in Q4 of 2018 (P<0.01) in all HF, 1.76% to 3.85% (P<0.01) for HFrEF, 2.22% to 3.21% (P<0.01) for HFpEF. (**Table 3**)

**Table 3:**
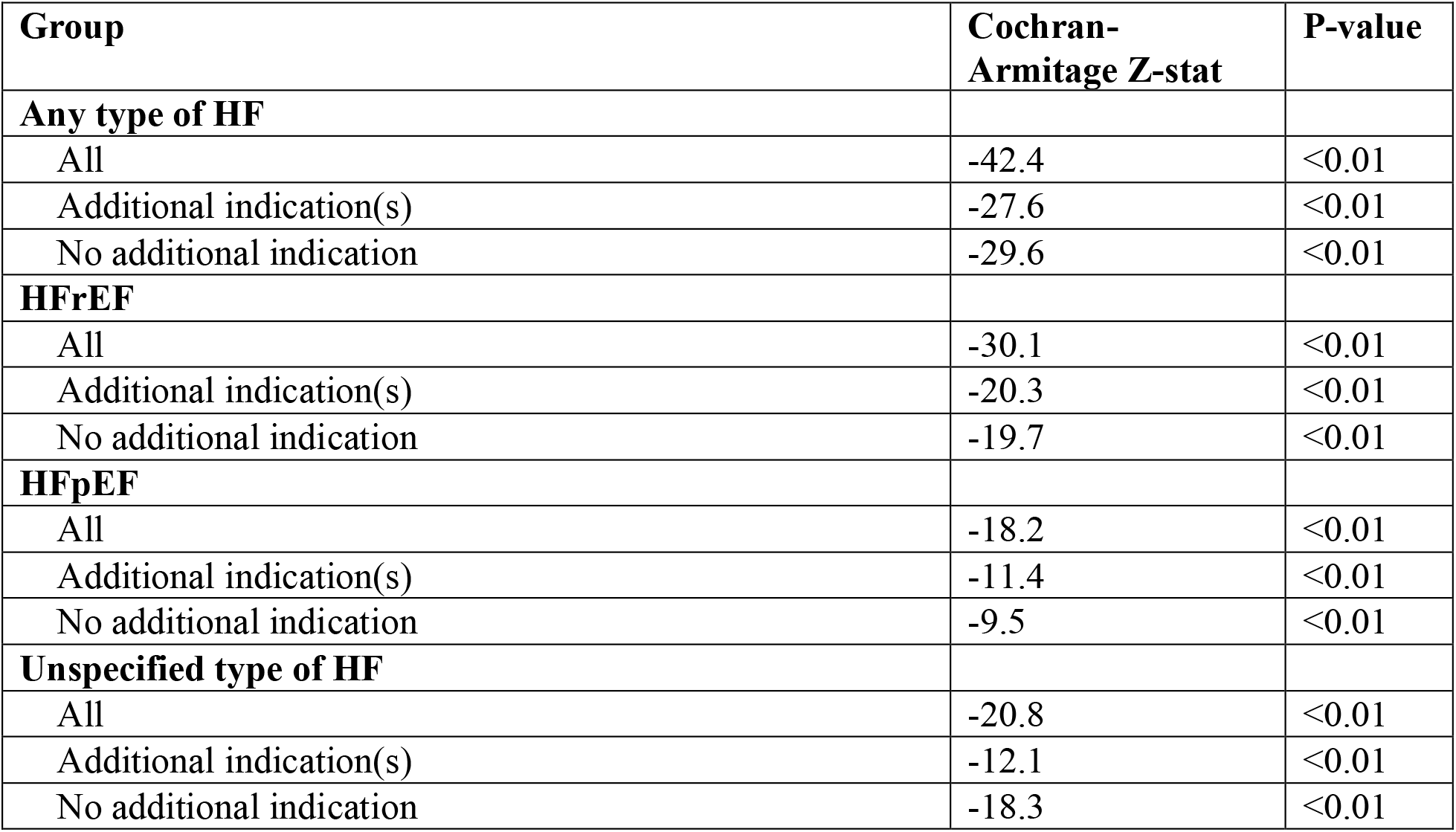
Cochran-Armitage Tests for Temporal Trends in Cardiac Rehabilitation Participation

| **Group** | **Cochran-Armitage Z-stat** | **P-value** |
| --- | --- | --- |
| **Any type of HF** |  |  |
| All | -42.4 | <0.01 |
| Additional indication(s) | -27.6 | <0.01 |
| No additional indication | -29.6 | <0.01 |
| **HFrEF** |  |  |
| All | -30.1 | <0.01 |
| Additional indication(s) | -20.3 | <0.01 |
| No additional indication | -19.7 | <0.01 |
| **HFpEF** |  |  |
| All | -18.2 | <0.01 |
| Additional indication(s) | -11.4 | <0.01 |
| No additional indication | -9.5 | <0.01 |
| **Unspecified type of HF** |  |  |
| All | -20.8 | <0.01 |
| Additional indication(s) | -12.1 | <0.01 |
| No additional indication | -18.3 | <0.01 |

**Figure 1.**
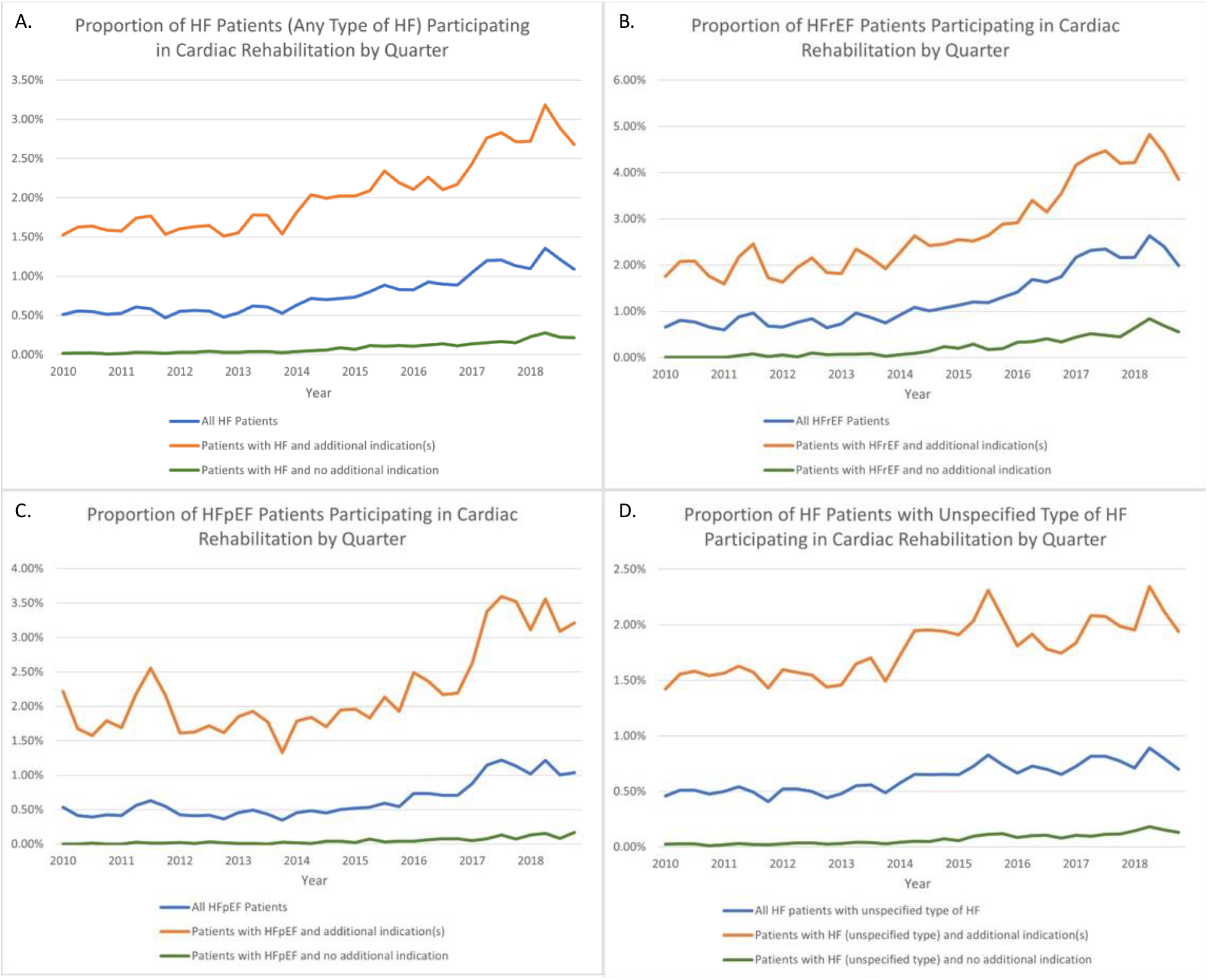
Percentage of patients who participated in CR during each quarter-year from 2010-2018. (A) patients with any type of HF, (B) patients with HFrEF, (C) patients with HFpEF, (D) patients with unspecified type of HF.

Payer source was a significant predictor of participation in CR (**Table 4**). Compared to patients with commercial insurance, patients who were covered by Medicare, Medicaid, or Medicare Advantage were significantly less likely to have participated in CR at least once from 2010 to 2018 ([Medicare; OR=0.697, P<0.001] [Medicaid; OR=0.563, P<0.001] [Medicare Advantage; OR=0.313, P<0.001]) Compared to patients with Medicare, patients with Medicaid were less likely to have participated in CR (OR 0.808, P<0.0001). and patients covered by Medicare Advantage were less likely to have participated in CR than patients covered by Medicaid (OR 0.556, P<0.0001). We assessed several clinical characteristics and their relationship with cardiac rehab participation in **table 4**. Patients who identify as non-white and older were less likely to participate in CR (Non-White; OR=0.850, [95%CI; 0.792-0.911], P<0.0001, Age; OR= 0.972, [95%CI: 0.970-0.974], P<0.0001). However male patients were more likely to participate (OR=1.435, [95%CI: 1.361-1.512], P<0.0001). In addition, patients with HFrEF were more likely to have participated in CR than patients with HFpEF (OR=1.380 [95% CI 1.266 – 1.504], P<0.0001). Patients who had at least one additional non-HF indication for CR were also more likely to have participated in CR=1.789 [95% CI1.696 – 1.888], P<0.0001). Out of individual comorbidities, coronary artery disease, history of a valve procedure, hypertension, hyperlipidemia, tobacco use, and cancer were positively associated with CR participation, whereas peripheral vascular disease, chronic obstructive pulmonary disease, chronic kidney disease, and diabetes mellitus were negatively associated with CR participation.

**Table 4:** Logistic Regression Analysis of Participation in Cardiac Rehabilitation from 2010-2018 by Payer Source, Adjusted for Demographic and Clinical Variables (n = 263,325)

| Predictor | Odds Ratio | 95% Confidence Interval |  | P-value |
| --- | --- | --- | --- | --- |
|  |  | Lower Bound | Upper Bound |  |
| Payer source: |  |  |  |  |
| Medicare vs. Commercial | 0.697 | 0.597 | 0.813 | <0.0001 |
| Medicaid vs. Commercial | 0.563 | 0.479 | 0.660 | <0.0001 |
| Medicare Advantage vs. Commercial | 0.313 | 0.260 | 0.377 | <0.0001 |
| Medicaid vs. Medicare | 0.808 | 0.743 | 0.879 | <0.0001 |
| Medicare Advantage vs. Medicare | 0.449 | 0.402 | 0.502 | <0.0001 |
| Medicare Advantage vs. Medicaid | 0.556 | 0.486 | 0.636 | <0.0001 |
| Other race vs. white | 0.850 | 0.792 | 0.911 | <0.0001 |
| Male sex vs. female sex | 1.435 | 1.361 | 1.512 | <0.0001 |
| Age (years) | 0.972 | 0.970 | 0.974 | <0.0001 |
| Type of heart failure: |  |  |  |  |
| HFrEF vs. unspecified | 1.610 | 1.511 | 1.715 | <0.0001 |
| HFpEF vs. unspecified | 1.167 | 1.085 | 1.255 | <0.0001 |
| HFrEF vs. HFpEF | 1.380 | 1.266 | 1.504 | <0.0001 |
| Presence of additional, non-HF indication(s) for CR | 1.789 | 1.696 | 1.888 | <0.0001 |
| Comorbidities: |  |  |  |  |
| Coronary artery disease | 9.376 | 8.435 | 10.422 | <0.0001 |
| History of valve procedure (surgery/TAVR) | 5.516 | 5.182 | 5.871 | <0.0001 |
| Atrial fibrillation | 1.017 | 0.965 | 1.073 | 0.5275 |
| Hypertension | 1.188 | 1.053 | 1.339 | 0.0051 |
| Hyperlipidemia | 2.981 | 2.686 | 3.307 | <0.0001 |
| Peripheral vascular disease | 0.914 | 0.865 | 0.966 | 0.0014 |
| Tobacco use | 1.338 | 1.269 | 1.411 | <0.0001 |
| Chronic obstructive pulmonary disease | 0.682 | 0.647 | 0.718 | <0.0001 |
| Cancer | 1.494 | 1.412 | 1.581 | <0.0001 |
| Chronic kidney disease | 0.857 | 0.814 | 0.903 | <0.0001 |
| Diabetes mellitus | 0.924 | 0.877 | 0.973 | 0.0027 |

## Discussion

Among insured patients with HF in Colorado within our study period of 2010-2018, CR participation has improved, however remains low. These data suggest CR participation is higher among patients with specific sociodemographic or comorbidity characteristics, including commercial insurance, male sex, white race, or having multiple indications for CR. This study highlights improvement in CR participation since the Medicaid expansion in 2014, however disparities in CR participation still exist.

Our finding of better CR participation and adherence among patients with commercial insurance can be compared to similar studies evaluating utilization of HF pharmacotherapy. This disparity between commercial insurance and Medicare or Medicaid insurances has been well documented across the medical field. One study by *Ozaki et al* demonstrated 4-fold more prescriptions for sacubitril/valsartan were dispensed in patients with commercial insurance compared to Medicare.^18^ This was despite a higher rate of prior authorization requirements.

These authors hypothesized that higher co-payment rates in patients with Medicare may contribute to poor utilization, as nearly 40% of patients with Medicare reported a co-payment of $40 or more.^18^ Moreover, a study by *Kini et al* demonstrated lower utilization of high-value cardiovascular disease testing among patients with Medicare and Medicaid compared to commercial insurance.^19^ A study by the *Medicaid Access Study Group* investigated the effect of insurance on access to outpatient clinic appointments by tasking research assistants with calling a variety of outpatient healthcare practices to make an appointment within two business days. The study found better access for patients with commercial insurance, and the most common reason for being unable to obtain an appointment was “Not accepting Medicaid”. Facilities that accepted Medicaid were less likely to have any appointments available.^20^ A similar phenomenon may be taking place with heart failure patients referred to CR, whereas many facilities may not accept Medicaid, and those that do may not have the resources to take on further patients. However further studies are needed to validate this hypothesis for CR.

Data supporting the use of CR in HF has been well established by multiple clinical trials and is reflected in the HF guidelines. ^3,5,6 21,22^ Despite this, our data show utilization of CR among patients with HF remains low and falls behind utilization among patients for other indications including percutaneous coronary intervention and acute coronary syndrome. ^10,23^ Furthermore, our study demonstrated patients who have multiple indications for CR but including HF still have low participation rates. This raises suspicion there is something specific about the HF population that may lead to poor CR participation rates.

Furthermore, Medicare reimbursement for CR for HFrEF was last updated in 2014 during our study period^12^ Yet our data show that overall participation in CR by patients with HF, even among those with HFrEF and/or *additional* indications for CR, is very low (<5%) as recently as 2018. Thus, advances are still needed to overcome remaining barriers to CR participation. These data echo those of Pandey et al., who showed similarly low overall CR participation rates (4.6%) and statistically significant but small absolute increases in 2014-2016 among Medicare beneficiaries.^17^ Our study demonstrates incremental improvement in CR participation among patients with HF from the time period of 2010 to 2018, regardless of insurance.

Many factors may potentially explain the very low observed CR participation rates among patients with HF. HF is a disease of aging – with multimorbidity and polypharmacy common among patients with HF. This is reflected in our data, which showed an average age of 74.3 years and high rates of cardiac and non-cardiac comorbidities. CR programs stated in a previous survey that patients with HF often declined to engage in CR because they were “simply too unwell to attend.”^24^ Patients with heart failure also tend to have a high prevalence of cognitive impairment,^25^ which can limit patients’ ability to travel and demands high caregiver engagement to allow patients to successfully participate. Indeed, transportation difficulty is a significant barrier to CR adherence for all eligible patients.^26^ There may also be residual, outdated concern that patients with HF cannot safely participate in CR.^27^ In addition, patients who have recently suffered a heart failure exacerbation often have profound skeletal muscle weakness, lack of balance, and/or poor mobility, and are inappropriate candidates for traditional CR, which typically focuses on moderate intensity aerobic activity.^28^ These patients require a more intensive, tailored approach, such as the protocol successfully tested in the REHAB-HF study.^29^ Furthermore, patients with HF often experience a heavy burden of symptoms, such as lack of energy, shortness of breath, and pain,^30,31^ which may limit their motivation to participate in CR and may cause CR staff to be wary of allowing patients to exercise effectively. All these factors likely contribute to the low CR participation rates observed in this study.

Despite the numerous barriers to improve CR participation among patients with HF, the significant increases in CR participation that we found, albeit small in magnitude, are also encouraging. For example, participation rates in patients with HFrEF tripled from 0.66% to 1.99% over eight years, with greater increases seen after 2014. The slightly higher increase in CR participation in patients with HFrEF compared to other types of HF suggests that Medicare reimbursement in the HFrEF population had a positive effect on CR participation. However, our data also suggest reimbursement is not a primary factor affecting CR participation, because participation increased among patients with types of HF that are not eligible for Medicare-reimbursed CR participation.

### Strengths and Limitations

Due to the comprehensive nature of our data source – the Colorado All Payers Claims Database – we were able to include all patients with a diagnosis of heart failure, regardless of payer source or site of care (i.e. inpatient vs. outpatient), in the state of Colorado over a long period of time (8 years) that spanned the change in reimbursement for CR for HFrEF patients. We also compared CR participation rates by type of heart failure and by presence of additional, non-HF indications for CR (e.g. coronary artery disease). All of these features build on existing large database analyses examining the problem of CR participation in patients with HF.^9,17^ Like all studies performed using claims data, our study has a lack of clinical nuance in defining the study population. For example, the type of heart failure (ischemic vs non-ischemic) for most patients in this study was unspecified. In addition, this study was limited to patients in Colorado, who may not be representative of the entire US population. Furthermore, data on only CR participation, not referral, were available, so it is unknown how many patients were referred but never attended a session. Finally, we did not differentiate between outpatient and inpatient claims in this study.

### Future Directions

Further studies of exercise-based rehabilitation interventions tailored to patients with HF are needed to increase the availability of programs that are effective and feasible given the unique features of the heart failure population. The REHAB-HF protocol was effective in improving physical function among older adults hospitalized for HF.^29^ How this protocol might be effectively rolled into existing clinical CR programs and reimbursed by all payers still needs to be established. In addition, more work is needed to build upon the REHAB-HF protocol to ensure exercise rehabilitation is effective and approachable for patients with HF who prefer home-based cardiac rehabilitation, and those who may need extra social or psychological support in managing their already high healthcare burden before they can successfully complete a cardiac rehabilitation program. Future study should also examine whether patients with HF do not participate in CR because of lack of referral, and if so, whether lack of referral is because of providers’ assumption that CR is inappropriate for the patient, patient disinterest, or both.

## Conclusion

This study shows that cardiac rehabilitation participation in patients with HF increased from 2010 to 2018. CR participation is highest in those with HFrEF, additional indications for CR, and commercial insurance, however overall rates of participation remain very low. These data suggest that although reimbursement for CR in patients with HF is important to augmenting CR participation, it is insufficient. More work is needed to develop and provide tailored, effective options for CR that would lead to broader participation within the heart failure population.

## Data Availability

Colorado All payer Claims Database

## Abbreviations

(CR): Cardiac rehabilitation
(HF): Heart failure
(HFpEF): Heart failure with preserved ejection fraction
(HFrEF): Heart failure with reduced ejection fraction.

## Acknowledgements

None

## Sources of funding

No sources of funding to acknowledge

## Disclosures

There are no industry relationships to disclose.

## Notes

### Competing Interest Statement

The authors have declared no competing interest.

### Clinical Trial

N/A

### Author Declarations

IRB of the Rocky Mountain Regional VA Medical Center

